# Simple model for Covid-19 epidemics – back-casting in China and forecasting in the US

**DOI:** 10.1101/2020.03.31.20049486

**Authors:** Slav W. Hermanowicz

## Abstract

In our previous work, we analyze, in near-real time, evolution of Covid-19 epidemic in China for the first 22 days of reliable data (up to February 6, 2020). In this work, we used the data for the whole 87 days (up to March 13, 2020) in China and the US data available till March 31 (day 70) for systematic evaluation of the logistic model to predict epidemic growth. We sequentially estimated sets of model parameters (maximum number of cases *K*, growth rate *r*, and half-time *t*_*0*_) and the epidemic “end time” *t*_*95*_ (defined as the time when the number of cases, predicted or actual, reached 95% of the maximum). The estimates of these parameters were done for sequences of reported cases growing daily (back-casting for China and forecasting for the US). In both countries, the estimates of *K* grew very much in time during the exponential and nearly exponential phases making longer term forecasting not reliable. For the US, the current estimate of the maximum number of cases *K* is about 265,000 but it is very likely that it will grow in the future. However, running estimates of the “end time” *t*_*95*_ were in a much smaller interval for China (60 – 70 days *vs*. the actual value of 67). For the US, the values estimated from the data sequences going back two weeks from now range from 70 to 80 days. If the behavior of the US epidemic is similar to the previous Chinese development, the **number of reported cases** could reach a **maximum around April 10 to 14**.

## Introduction

In our previous recent work (Hermanowicz 2020), we used a simple logistic model to analyze the evolution of data on Covid-19 cases as reported in mainland China by the National Health Commission of the People’s Republic of China (NHC 2020). This initial analysis was done in three phases in near-real time (up to Jan. 30, Feb. 3, and Feb 6). The analysis resulted in a sequence of continually updated forecasts of the epidemic dynamics. Although the predicted maximum numbers of cases increased as new data became available, they systematically underestimated final reported numbers (even without accounting for a major change in reporting criteria in China on February 12 resulting in a large surge of new reported cases). However, the successive **predicted dynamics** of the epidemic were **remarkably close** to the final real-world outcome. We explore this issue further in this work where we use the full reported dataset for mainland China for **systematic back-casting** of the epidemic.

At the same time, when this work is being conducted, in the US we are experiencing a fast developing Covid-19 epidemic that in some aspects is similar to China but with a delay. We use the experience from our analysis of the situation in China to **forecast** further evolution of coronavirus cases in the United States. Obviously, our predictions are based on the data currently available. These predictions do not account for any possible other secondary sources of infection, changes of diagnostics or reporting, or virus mutation.

Modelling the epidemics and infection dynamics is very important and numerous results have been recently reported for worldwide Covid-19 outbreak (Chen, Rui et al. 2020, Chen, Cheng et al. 2020, Hong, He et al. 2020, Hui, Azhar et al. 2020, Imai, Cori et al. 2020, Imai, Dorigatti et al. 2020, Li, Guan et al. 2020, Linton, Kobayashi et al. 2020, Liu, Hu et al. 2020, Liu, Hewings et al. 2020, Roosa, Lee et al. 2020, Shen, Peng et al. 2020, Wu, Leung et al. 2020, Zhang and Wang 2020, Zhao, Lin et al. 2020, Zhou, Hong et al. 2020, Ziff and Ziff 2020). Many reported models are complicated, incorporate tentative assumptions and need parameters estimates that are not reliable as underscored by Fong and coworkers (Fong, Li et al. 2020).

In this work, we present the results of fitting a very simple logistic model to the available data and a forecast of new infections. In contrast to other models, the logistic model does not include any external assumptions and is derived completely from available data.

### Logistic Model and its Application

The logistic model is one of the simplest used in population dynamics and has been used specifically for epidemics for a long time (Bailey 1950, Cockburn 1960, Mansfield and Hensley 1960, Jowett, Browning et al. 1974, Waggoner and Aylor 2000, Koopman 2004, Bangert, Molyneux et al. 2017). In our earlier analysis of the developing epidemic in China, we used a discrete version of the model due to its uncomplicated structure and easy calculations.

In discrete time, more appropriate to daily reported infection cases, the logistic model becomes

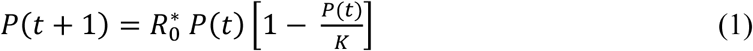

where *P*(*t*) and *P*(*t*+*1*) are populations (cases) on consecutive days, *R*_*0*_^*^ is the growth rate (basic reproduction number in epidemiology) at the beginning of the logistic growth, and *K* is the limiting population (maximum cases).

However, expressing the growth of population *P* in continuous time *t* allows to formulate the model as an ordinary first-order differential equation describing dynamic evolution of the population *P* (in our case the number of infected individuals) being controlled by the growth rate *r* and maximum cases *K* due to limited growth. In continuous time *t*, the change of *P* is

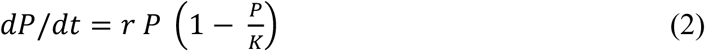

Initially, the growth of *P* is close to exponential since the term (1− *P*/*K*)is almost one. When *P* becomes larger (commensurate with *K*) the growth rate slows down with

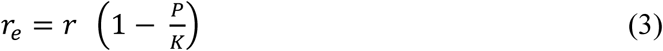

becoming an effective instantaneous growth rate.

The solution to Eq. (2) is the well-known sigmoidal function (logistic function)

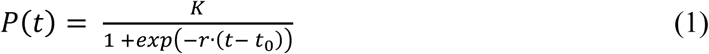

where *t*_*0*_ is the time when the population reaches one-half of the maximum value *P*(*t*_*0*_) *= 1/2 K*. Using a differential version of the model is more convenient since a closed-form solution exists and allows for direct estimation of three model parameters: *K, r*, and *t*_*0*_. The logistic model may be adequate for the analysis of mainland China and the US as a whole since at this time each country can be treated as a unit where a vast majority of cases occurred without any ***significant*** “import” or “export” of cases.

### Data Analysis

For China, we used data reported daily by the National Health Commission of the People’s Republic of China (NHC 2020) up to March 13, 2020 (day 87 from the outbreak) when only 11 new cases were reported – less than 2·10^−4^ of total case number, effectively ending the epidemic on the national level. There is a considerable controversy as to the exact date of the outbreak with most reports pointing to mid-December (Li, Guan et al. 2020, Wang, Horby et al. 2020) while one analysis suggest multiple sources of original infection (Nishiura, Jung et al. 2020). Initially, the outbreak was not recognized and number of ***confirmed*** cases is not fully known (Wu, Hao et al. 2020). In our analysis, we adopted December 17, 2019 as the best estimate of the outbreak following the work of Zhang and co-workers (Zhang and Wang 2020, Zhang and Wang 2020). In addition, in the initial stages of the epidemic the reported numbers of cases may severely underestimate the actual numbers due to asymptomatic carriers (Zhao, Musa et al. 2020). More accurate numbers can be only estimated after the epidemic (Wu and McGoogan 2020).

The data from China are show in Figure 1 and in the Appendix. As seen in this figure, the cumulative number of cases grew in a sigmoidal fashion. On February 12, 2020 (day 57) the reporting criteria were changed resulting in a one-day increase of about 15,000 cases. We analyzed the subsequent data with this jump and also ignoring it. The logistic model turned out to be quite robust. While the numerical estimates of the maximum number of cases *K* was significantly affected by the inclusion or exclusion the jump, the dynamics of the model was much less impacted (see further discussion). In this work, we decided to report the results with the jump as parsing data of new and cumulative cases past day 57 is very unreliable.

**Figure 1.**
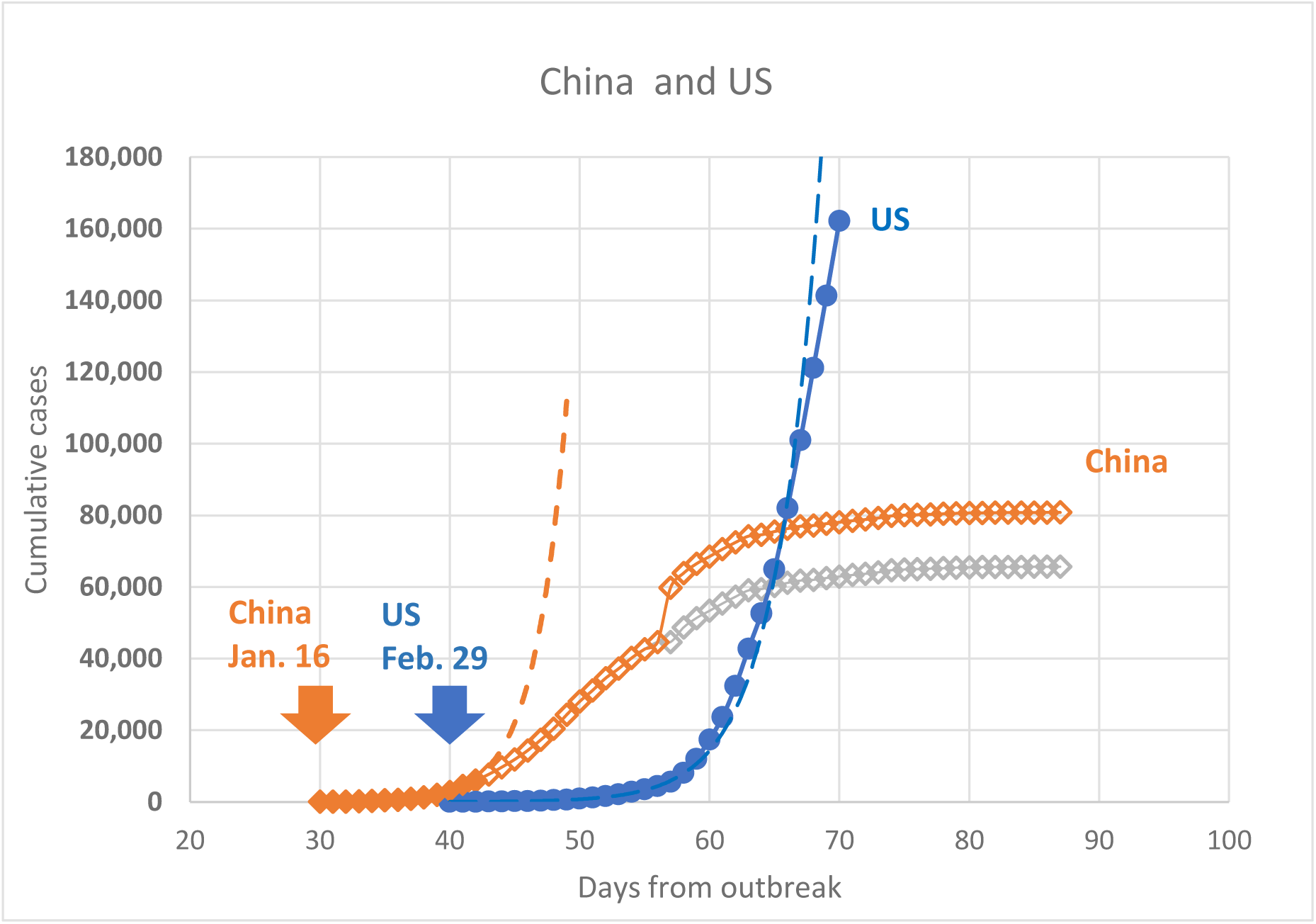
Evolution of reported cases in China and the US. (Solid points show data used to fit exponential functions – dashed lines)

In the US, there are numerous agencies tracking the Covid-19 cases at different administrative levels in accordance with multi-tiered structure of the US government. For this reason, we relied on web-based data aggregators (mostly at state level) following daily updates (Covid-19 2020).

We used January 21, 2020 as the start of the epidemic in the US (day 1) since on that day the first non-repatriated Covid-19 case was reported. It should be noted that like in China the number of cases in the initial stages is underreported due to the same factors (asymptomatic carriers, initial administrative confusion, limited testing). Despite these problems, the data used in this work are the best currently available. The current evolution of the epidemic in the US is presented in Figure 1 and in the Appendix. Despite smaller population, the number of cases in the US at the time of writing already exceeds that of China. Reasons for such higher numbers are outside the scope of this analysis.

Both, China and the US data are also plotted in the semi-log format (??) to underscore exponential growth of the epidemic in the initial stages – straight line on these plots. In China, exponential growth occurred from day 30 till day 42 as shown in ?? and ?? by the dashed lines. After day 42 (January 28), the number of cases still increased but the rate of growth was becoming lower and the line representing the cumulative cases deviated from the exponential curve. This feature was also clearly recognized in a previous report (Zhao, Lin et al. 2020).

In the US, exponential growth occurred for a longer period and only very recently (March 27 – 30) **perhaps** starts to deviate from the exponential curve.

## Model Estimates and Results

For each dataset of reported cumulative case numbers (China and the US), we estimated three parameters of the logistic model (maximum case number *K*, growth rate *r*, half-time *t*_*0*_) fitting model predictions to the data. We use a custom nonlinear curve fitting procedure employing the Levenberg-Marquardt method for minimization of the residual sum of squares. Similarly to our previous work (Hermanowicz 2020), we estimated model parameters sequentially from datasets growing day after day.

For China, the first dataset contained 5 days from day 38 through 42. The next estimate used 6 days from day 38 through 43. This process was repeated until day 87 when the entire China dataset was used. For the US, the first dataset contained also 5 days from day 43 through 47. The last day of the available sequence for the US was day 70 (March 30, 2020). All resulting estimates are show in the Appendix.

In case of China, where the epidemic growth has essentially ended, the sequential estimation process (back-casting) simulated near-real time analysis of the dynamics. In the US, where we are still in the substantial growth phase, the sequential estimates are indeed performed in near-real time. In addition to three model parameters, for each day we also estimated predicted time for the epidemic to end. For this purpose. we chose arbitrarily time when the predicted number of cases reach 95% of the predicted maximum *K*. This time, *t*_*95*_ was calculated from Eq. (4) by setting *P*(*t*) *= 0*.*95 K* and is also shown in the Appendix.

### China

Figure 3 shows the development of sequential estimates of the maximum predicted cases *K* for China and Figure 4 presents the corresponding “ending times” *t*_*95*_. As reported earlier (Hermanowicz 2020) for actual near-real time analysis, estimates of predicted maximum cases *K* depended very heavily on the length of the dataset used for estimation. As examples, Figure 5 shows a few logistic curves corresponding to selected model parameter estimates on specific days for China. As seen in this figure, the initial estimates (close to the exponential phase) were below 20,000 but they grew to about 100,000 as more data became available and was used for estimate refinement. Obviously, the estimate of *K* obtained from the full dataset (up to day 87) matches closely the actual reported number of cases (80,780 *vs*. 80,807) but it should be noted that the estimates of *K* from day 65 (more than 20 days in advance) converged very closely to the actual maximum.

**Figure 2.**
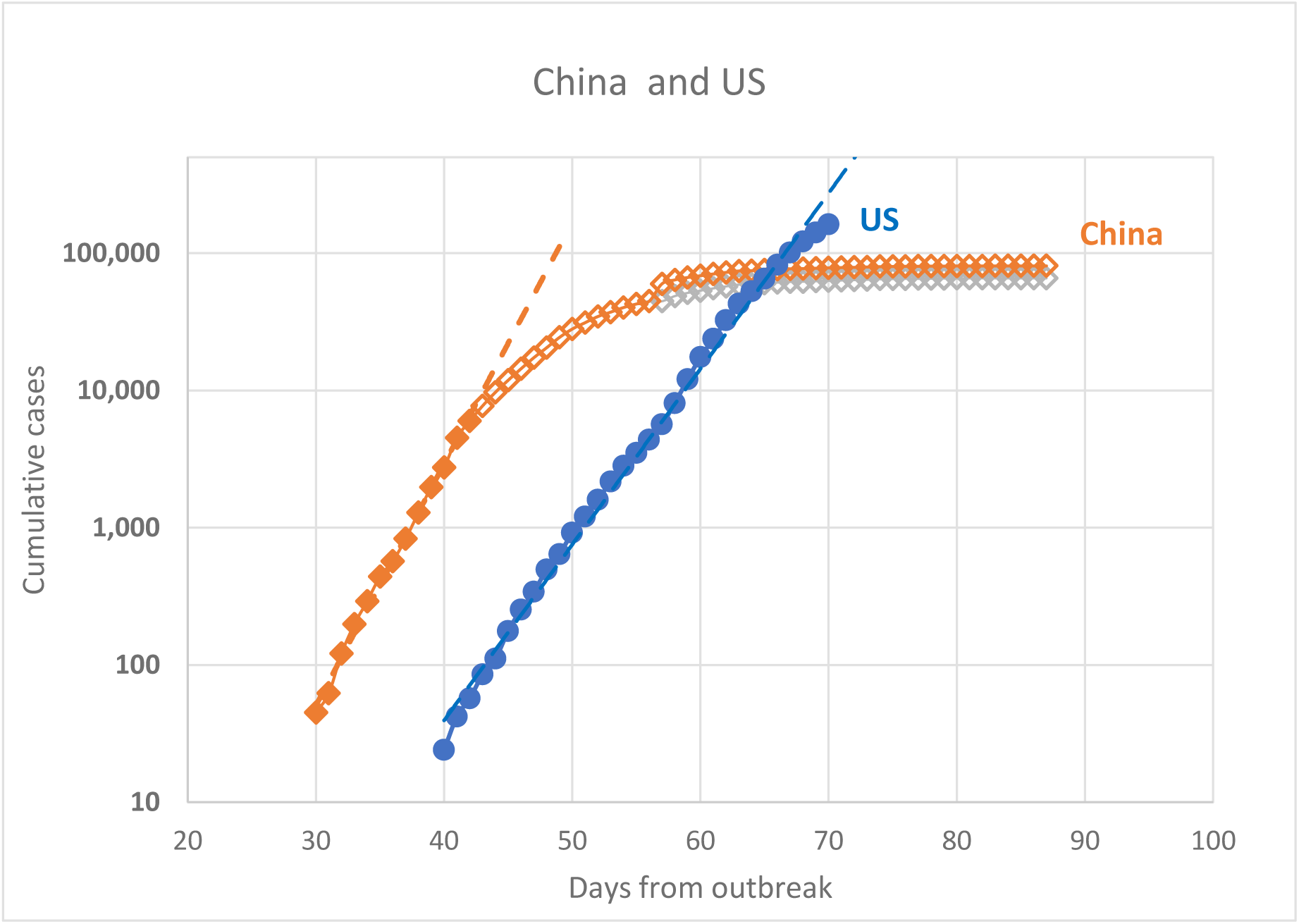
Exponential growth of epidemics (see Figure 1 for notes)

**Figure 3.**
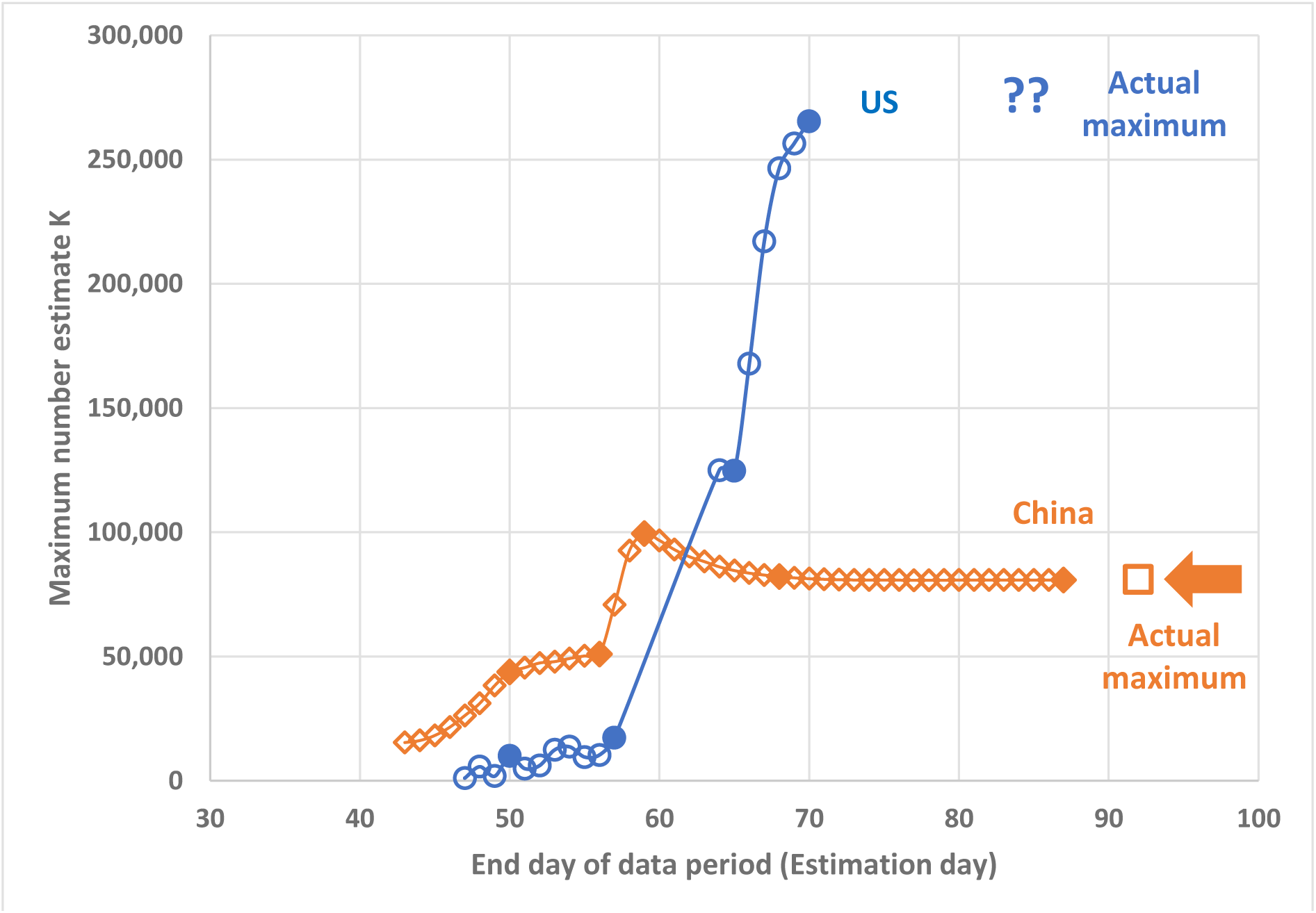
Sequential estimates of K (solid data points show days for which logistic model predictions are shown in Figure 5 for China and Figure 6 for the US)

**Figure 4.**
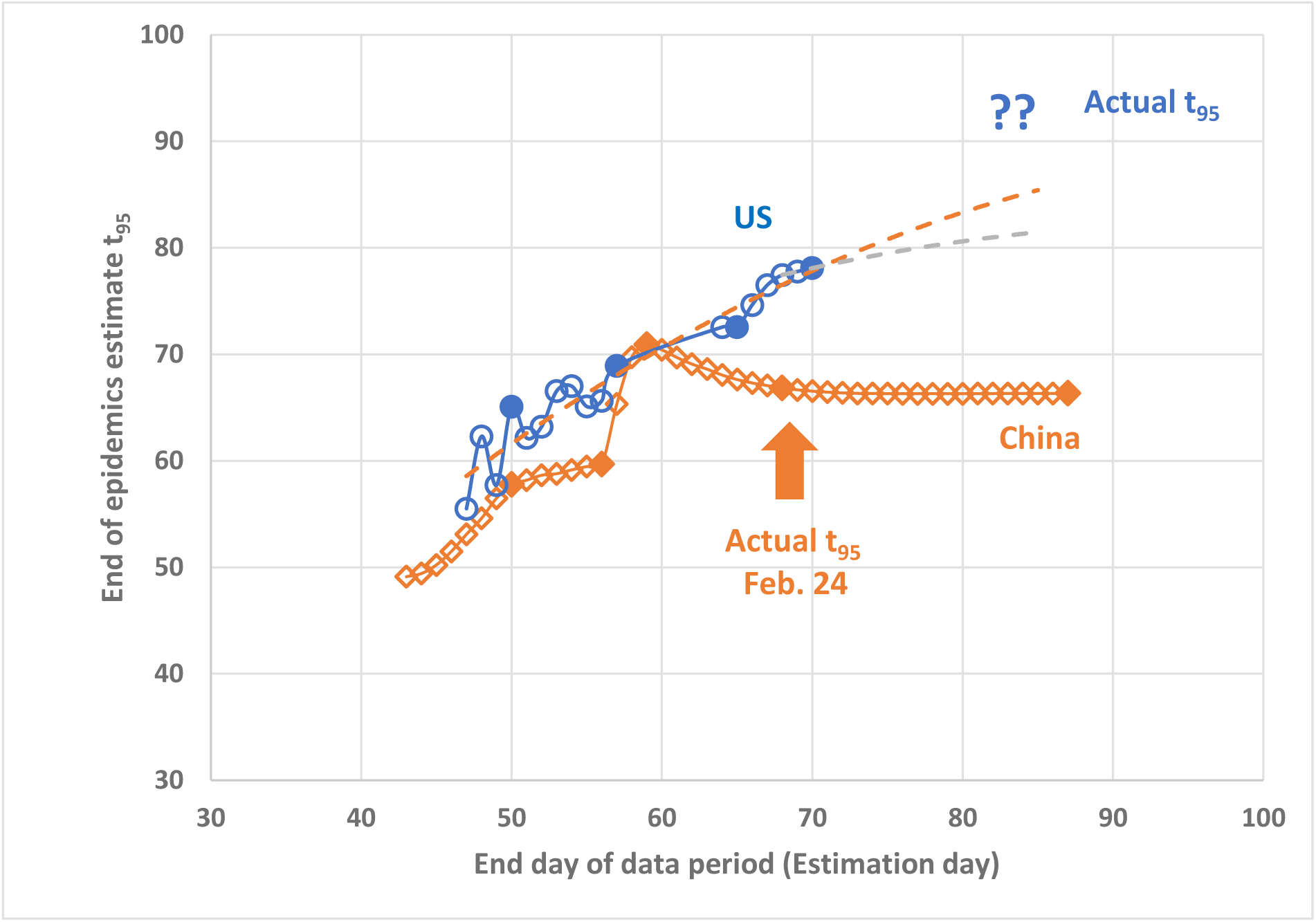
Sequential estimates of “end time” t_95_ (see notes in Figure 3)

**Figure 5.**
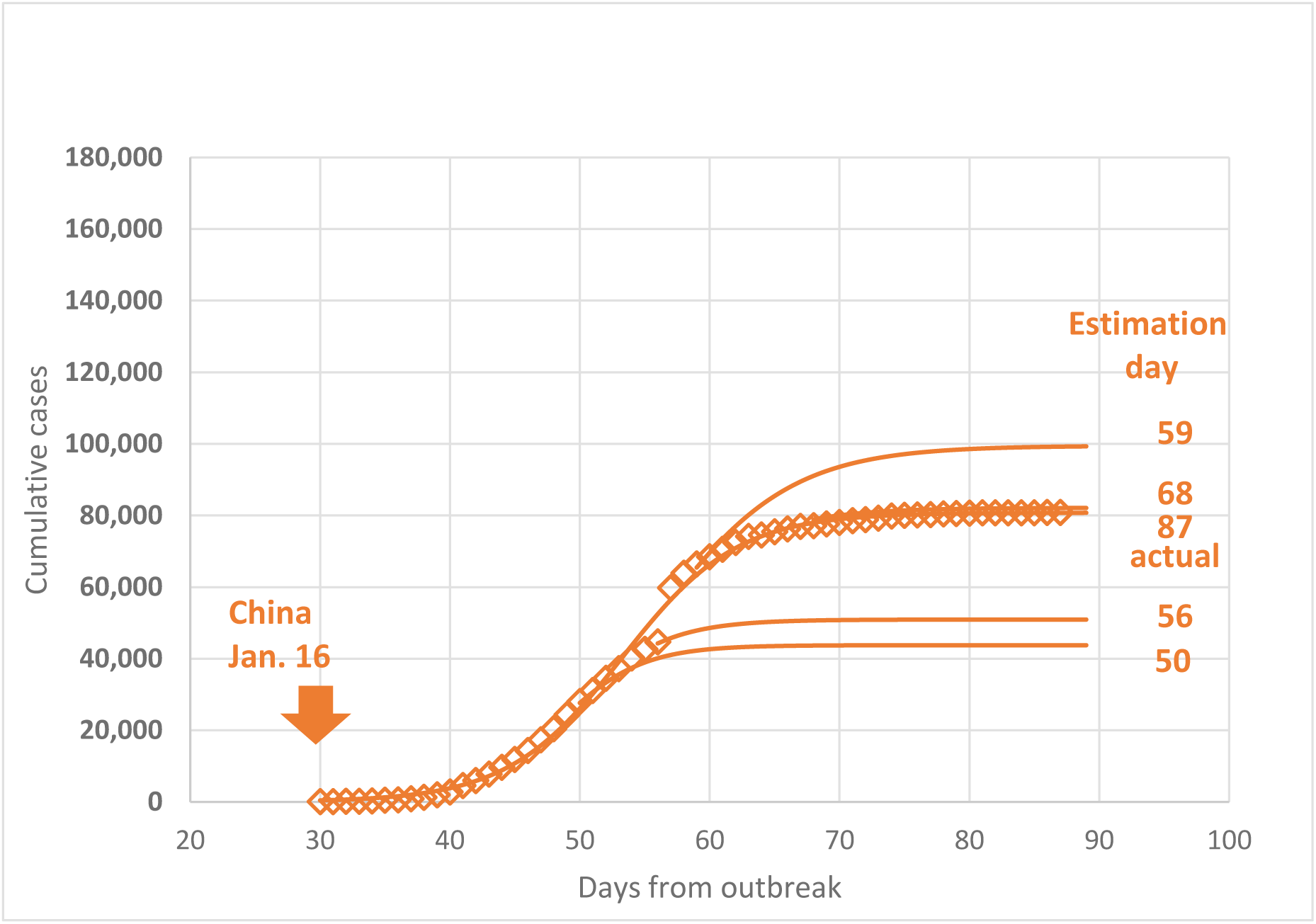
Examples of model predictions for China for selected parameter estimates on specific days

Due to a change in reporting criteria in China on day 57 (February 12), there was a major increase in the number of reported cases that could not be incorporated in the model. As mentioned before, we decided to use all data (with the jump) in any subsequent analysis. The immediate result was a very large increase in the estimated maximum cases *K* (Figure 3), approximately doubling it from about 50,000 on day 56 to about 99,400 on day 59. This big increase underscores again the sensitivity of the logistic model to data quality. However, the model is also robust in a longer term as the *K* estimates quickly converged toward their ultimate values.

Similarly, there was also a significant jump in the estimates of *t*_*95*_ (see Figure 4) due to the jump in the case numbers. However, it is remarkable that the estimates of the “end time” *t*_*95*_ were much more constrained and much closer to the actual value. Even as early as three weeks before the end of the epidemic the estimates of the “end time” were between 60 and 70 days, very close to the final value of 67 days.

### United States

Unlike China, the epidemic in the US is still at the growth phase, perhaps deviating slightly from the exponential growth. Thus, the available dataset is much smaller and the logistic model estimates are burdened with much larger uncertainty. Currently available estimates of the maximum cases *K* are also shown in Figure 3. They exhibit very large variations increasing from approximately 1,100 on day 47 to approximately 265,000 on day 70 without any sign of leveling off. This large variation is not unexpected since the nearly exponential growth does not contain sufficient information on the actual maximum. In other words, the derivative *dP*/*dt* in Eq. (2) is dominated by the term *r P* while (*1*-*P*/*K*) ≈ *1*. This behavior is also seen in the examples of predicted logistic curves for selected sets of estimated parameters *K, r, t*_*0*_ (Figure 6).

**Figure 6.**
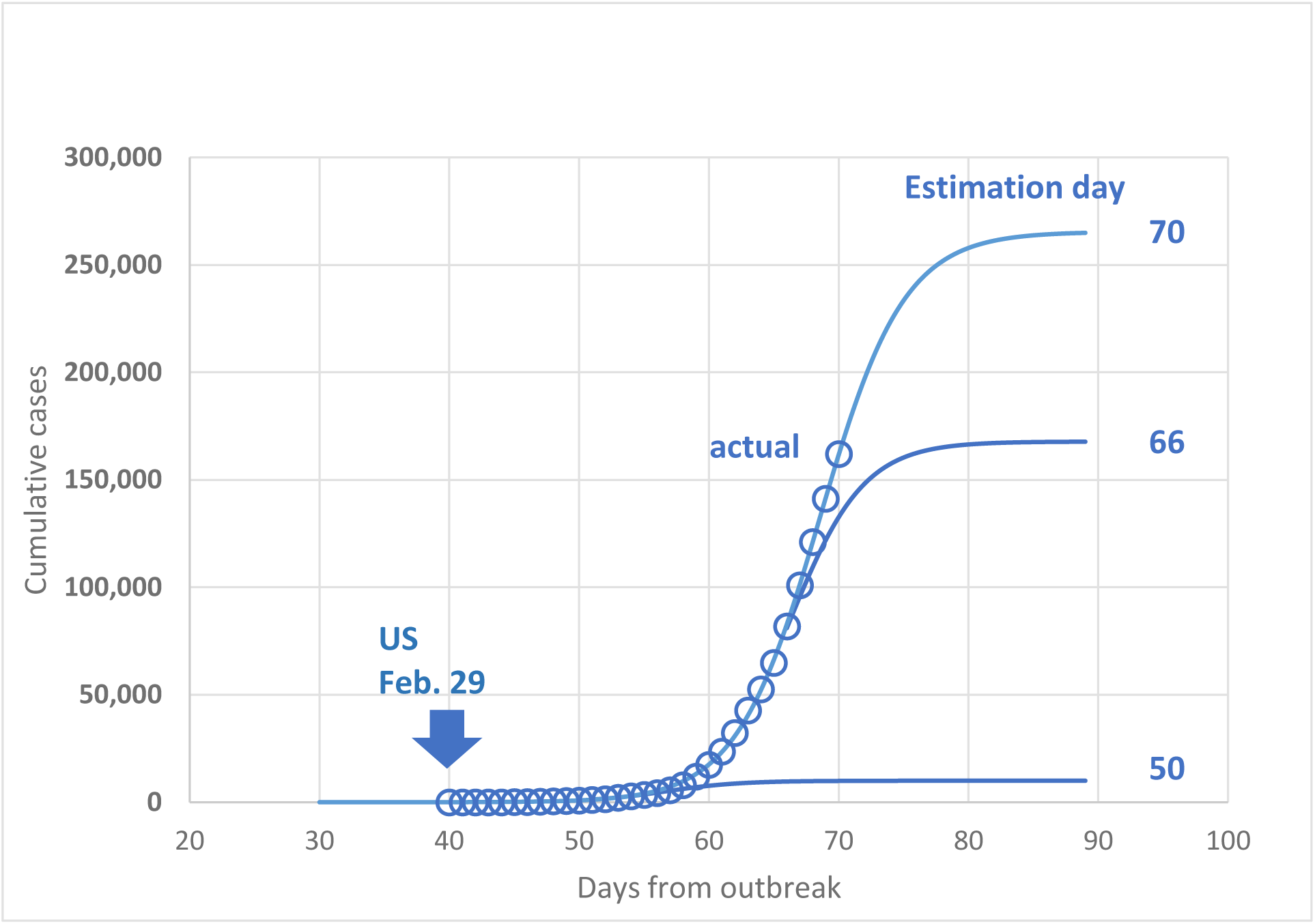
Examples of model predictions for the US for selected parameter estimates on specific days

The sequential estimates of the “end time” *t*_*95*_ are plotted in Figure 4. They also tend to increase in time with the increasing number of reported cases but unlike the estimates of *K* (and similarly to the Chinese case) it varies in a much smaller interval – 70 to 80 days in the past two weeks, If the behavior of the US epidemic is similar to the Chinese case, we could expect further leveling off of *t*_*95*_ at slightly above 80 days. If this **bold prediction** will hold, we could see the end of the epidemic growth around 80 – 85 days after its origin, around April 10 to 14. Of course, the end here is defined as the cessation of new cases and not the complete recovery of the infected patients.

## Data Availability

data in the paper

## Declarations

### Ethics approval and consent to participate

The ethical approval or individual consent was not applicable.

### Availability of data and materials

All data and materials used in this work were publicly available.

### Consent for publication

Not applicable.

### Disclaimer

The funding agencies had no role in the design and conduct of the study; collection, management, analysis, and interpretation of the data; preparation, review, or approval of the manuscript; or decision to submit the manuscript for publication.

## Appendix

**Table A- 1.** Number of cases in China

| Date | Day since outbreak | Cumulative Cases |
| --- | --- | --- |
| January 16, 2020 | 30 | 45 |
| January 17, 2020 | 31 | 62 |
| January 18, 2020 | 32 | 121 |
| January 19, 2020 | 33 | 198 |
| January 20, 2020 | 34 | 291 |
| January 21, 2020 | 35 | 440 |
| January 22, 2020 | 36 | 571 |
| January 23, 2020 | 37 | 830 |
| January 24, 2020 | 38 | 1,287 |
| January 25, 2020 | 39 | 1,975 |
| January 26, 2020 | 40 | 2,744 |
| January 27, 2020 | 41 | 4,515 |
| January 28, 2020 | 42 | 5,974 |
| January 29, 2020 | 43 | 7,711 |
| January 30, 2020 | 44 | 9,692 |
| January 31, 2020 | 45 | 11,860 |
| February 1, 2020 | 46 | 14,380 |
| February 2, 2020 | 47 | 17,307 |
| February 3, 2020 | 48 | 20,467 |
| February 4, 2020 | 49 | 24,324 |
| February 5, 2020 | 50 | 28,018 |
| February 6, 2020 | 51 | 31,161 |
| February 7, 2020 | 52 | 34,546 |
| February 8, 2020 | 53 | 37,198 |
| February 9, 2020 | 54 | 40,171 |
| February 10, 2020 | 55 | 42,638 |
| February 11, 2020 | 56 | 44,653 |
| February 12, 2020 | 57 | 59,804 |
| February 13, 2020 | 58 | 63,851 |
| February 14, 2020 | 59 | 66,492 |
| February 15, 2020 | 60 | 68,500 |
| February 16, 2020 | 61 | 70,548 |
| February 17, 2020 | 62 | 72,436 |
| February 18, 2020 | 63 | 74,185 |
| February 19, 2020 | 64 | 74,576 |
| February 20, 2020 | 65 | 75,465 |
| February 21, 2020 | 66 | 76,288 |
| February 22, 2020 | 67 | 76,936 |
| February 23, 2020 | 68 | 77,150 |
| February 24, 2020 | 69 | 77,658 |
| February 25, 2020 | 70 | 78,064 |
| February 26, 2020 | 71 | 78,494 |
| February 27, 2020 | 72 | 78,824 |
| February 28, 2020 | 73 | 79,251 |
| February 29, 2020 | 74 | 79,824 |
| March 1, 2020 | 75 | 80,026 |
| March 2, 2020 | 76 | 80,151 |
| March 3, 2020 | 77 | 80,270 |
| March 4, 2020 | 78 | 80,404 |
| March 5, 2020 | 79 | 80,547 |
| March 6, 2020 | 80 | 80,646 |
| March 7, 2020 | 81 | 80,690 |
| March 8, 2020 | 82 | 80,730 |
| March 9, 2020 | 83 | 80,749 |
| March 10, 2020 | 84 | 80,773 |
| March 11, 2020 | 85 | 80,788 |
| March 12, 2020 | 86 | 80,796 |
| March 13, 2020 | 87 | 80,807 |

**Table A- 2.**
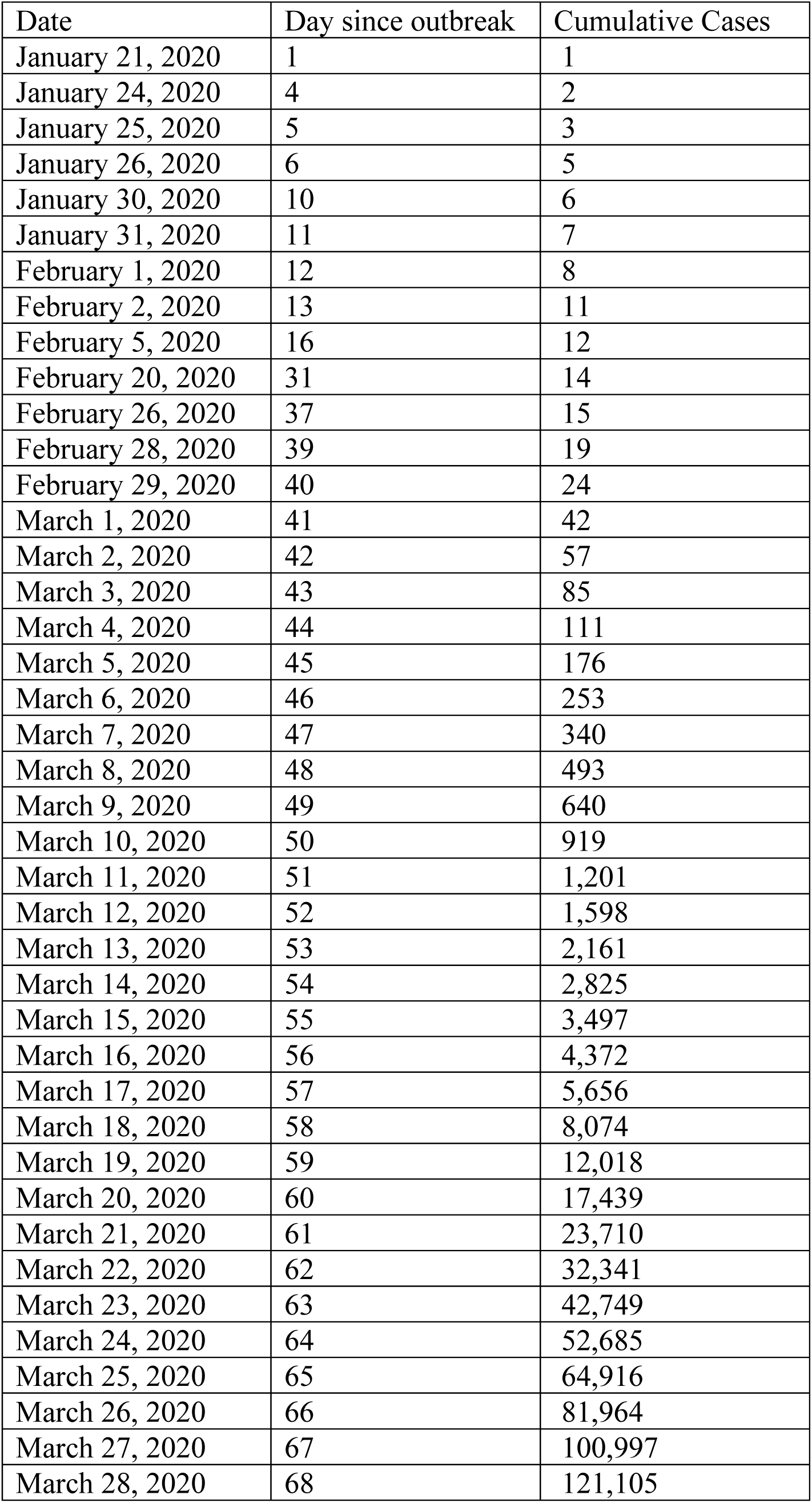
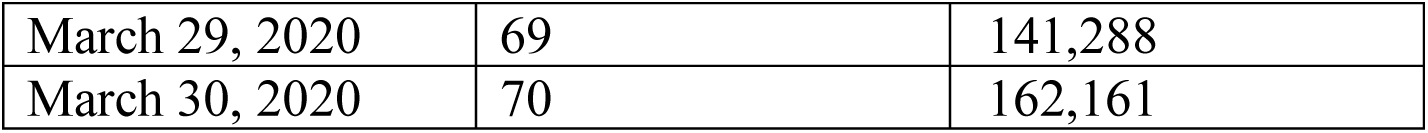
Number of cases in the US

**Table A- 3.** Sequential estimates of logistic model for China

| End day | Maximum $K$ | Rate $r$ | Mid-time $t_0$ | End time $t_{95}$ |
| --- | --- | --- | --- | --- |
| 42 | 2.257E+04 | 0.445 | 44.3 | 50.9 |
| 43 | 1.540E+04 | 0.479 | 43.0 | 49.1 |
| 44 | 1.620E+04 | 0.471 | 43.2 | 49.4 |
| 45 | 1.824E+04 | 0.448 | 43.7 | 50.2 |
| 46 | 2.157E+04 | 0.416 | 44.4 | 51.5 |
| 47 | 2.621E+04 | 0.382 | 45.4 | 53.1 |
| 48 | 3.116E+04 | 0.355 | 46.3 | 54.6 |
| 49 | 3.829E+04 | 0.326 | 47.5 | 56.5 |
| 50 | 4.377E+04 | 0.31 | 48.3 | 57.8 |
| 51 | 4.552E+04 | 0.304 | 48.5 | 58.2 |
| 52 | 4.740E+04 | 0.299 | 48.8 | 58.6 |
| 53 | 4.796E+04 | 0.297 | 48.9 | 58.8 |
| 54 | 4.921E+04 | 0.292 | 49.1 | 59.1 |
| 55 | 5.024E+04 | 0.288 | 49.2 | 59.5 |
| 56 | 5.093E+04 | 0.284 | 49.3 | 59.7 |
| 57 | 7.080E+04 | 0.226 | 52.3 | 65.3 |
| 58 | 9.266E+04 | 0.198 | 54.9 | 69.7 |
| 59 | 9.947E+04 | 0.192 | 55.6 | 70.9 |
| 60 | 9.672E+04 | 0.195 | 55.3 | 70.4 |
| 61 | 9.304E+04 | 0.199 | 54.9 | 69.7 |
| 62 | 9.017E+04 | 0.203 | 54.6 | 69.1 |
| 63 | 8.820E+04 | 0.206 | 54.3 | 68.6 |
| 64 | 8.610E+04 | 0.21 | 54.0 | 68.1 |
| 65 | 8.461E+04 | 0.213 | 53.8 | 67.6 |
| 66 | 8.356E+04 | 0.216 | 53.7 | 67.3 |
| 67 | 8.280E+04 | 0.218 | 53.6 | 67.1 |
| 68 | 8.214E+04 | 0.22 | 53.5 | 66.8 |
| 69 | 8.167E+04 | 0.222 | 53.4 | 66.7 |
| 70 | 8.132E+04 | 0.223 | 53.4 | 66.5 |
| 71 | 8.108E+04 | 0.224 | 53.3 | 66.4 |
| 72 | 8.091E+04 | 0.225 | 53.3 | 66.4 |
| 73 | 8.080E+04 | 0.226 | 53.3 | 66.3 |
| 74 | 8.077E+04 | 0.226 | 53.3 | 66.3 |
| 75 | 8.075E+04 | 0.226 | 53.3 | 66.3 |
| 76 | 8.074E+04 | 0.226 | 53.3 | 66.3 |
| 77 | 8.073E+04 | 0.226 | 53.3 | 66.3 |
| 78 | 8.073E+04 | 0.226 | 53.3 | 66.3 |
| 79 | 8.073E+04 | 0.226 | 53.3 | 66.3 |
| 80 | 8.074E+04 | 0.226 | 53.3 | 66.3 |
| 81 | 8.075E+04 | 0.226 | 53.3 | 66.3 |
| 82 | 8.076E+04 | 0.226 | 53.3 | 66.3 |
| 83 | 8.076E+04 | 0.226 | 53.3 | 66.3 |
| 84 | 8.077E+04 | 0.226 | 53.3 | 66.3 |
| 85 | 8.077E+04 | 0.226 | 53.3 | 66.3 |
| 86 | 8.078E+04 | 0.226 | 53.3 | 66.3 |
| 87 | 8.078E+04 | 0.226 | 53.3 | 66.3 |

Note: Each row contains estimates of the logistic model $K$ , $r$ , $t_0$ and the 95% end time $t_{95}$ estimated from recorded number of cases for the period starting on day 38 and ending on the day in Column 1.

**Table A- 4.** Sequential estimates of logistic model for the US

| End day | Maximum $K$ | Rate $r$ | Mid-time $t_0$ | End time $t_{95}$ |
| --- | --- | --- | --- | --- |
| 47 | 1.074E+03 | 0.436 | 48.8 | 55.5 |
| 48 | 5.659E+03 | 0.371 | 54.4 | 62.3 |
| 49 | 1.851E+03 | 0.41 | 50.5 | 57.7 |
| 50 | 1.000E+04 | 0.348 | 56.6 | 65.1 |
| 51 | 4.923E+03 | 0.365 | 54.1 | 62.1 |
| 52 | 6.144E+03 | 0.356 | 54.9 | 63.2 |
| 53 | 1.243E+04 | 0.333 | 57.7 | 66.6 |
| 54 | 1.368E+04 | 0.33 | 58.1 | 67.0 |
| 55 | 9.575E+03 | 0.345 | 56.6 | 65.1 |
| 56 | 1.042E+04 | 0.34 | 57.0 | 65.6 |
| 57 | 1.729E+04 | 0.309 | 59.4 | 68.9 |
| 64 | 1.250E+05 | 0.38 | 64.8 | 72.6 |
| 65 | 1.248E+05 | 0.38 | 64.8 | 72.5 |
| 66 | 1.678E+05 | 0.349 | 66.2 | 74.6 |
| 67 | 2.170E+05 | 0.327 | 67.5 | 76.5 |
| 68 | 2.465E+05 | 0.316 | 68.1 | 77.5 |
| 69 | 2.565E+05 | 0.313 | 68.4 | 77.8 |
| 70 | 2.654E+05 | 0.309 | 68.6 | 78.1 |

Note: Each row contains estimates of the logistic model $K$ , $r$ , $t_0$ and the 95% end time $t_{95}$ estimated from recorded number of cases for the period starting on day 43 and ending on the day in Column 1.

## References

Bailey, N. T. J. (1950). “A SIMPLE STOCHASTIC EPIDEMIC.” Biometrika 37(3-4): 193–202.

Bangert, M., D. H. Molyneux, S. W. Lindsay, C. Fitzpatrick and D. Engels (2017). “The cross-cutting contribution of the end of neglected tropical diseases to the sustainable development goals.” Infectious Diseases of Poverty 6.

Chen, T., J. Rui, Q. Wang, Z. Zhao, J.-A. Cui and L. Yin (2020) “A mathematical model for simulating the transmission of Wuhan novel Coronavirus.” bioRxiv, 2020.2001.2019.911669 DOI: 10.1101/2020.01.19.911669.

Chen, Y., J. Cheng, Y. Jiang and K. J. Liu (2020). “A time delay dynamic system with external source for the local outbreak of 2019-nCoV.” Applicable Analysis.

Cockburn, T. A. (1960). “EPIDEMIC CRISIS IN EAST PAKISTAN - APRIL-JULY, 1958.” Public Health Reports 75(1): 26–36.

Covid-19. (2020). “Template:2019–20 coronavirus pandemic data/United States medical cases.” Retrieved March 30. 2020, 2020, from https://en.wikipedia.org/wiki/Template:2019%E2%80%9320_coronavirus_pandemic_data/United_States_medical_cases.

Fong, S. J., G. Li, N. Dey, R. G. Crespo and E. Herrera-Viedma (2020). “Finding an Accurate Early Forecasting Model from Small Dataset: A Case of 2019-nCoV Novel Coronavirus Outbreak.” International Journal of Interactive Multimedia and Artificial Intelligence 6(1): 132–140.

Hermanowicz, S. W. (2020) “Forecasting the Wuhan coronavirus (2019-nCoV) epidemics using a simple (simplistic) model - update (Feb. 8, 2020).” medRxiv, 2020.2002.2004.20020461 DOI: 10.1101/2020.02.04.20020461.

Hong, N., J. He, Y. Ma, H. Jiang, L. Han, L. Su, W. Zhu and Y. Long (2020) “Evaluating the secondary transmission pattern and epidemic prediction of the COVID-19 in metropolitan areas of China.” medRxiv, 2020.2003.2006.20032177 DOI: 10.1101/2020.03.06.20032177.

Hui, D. S., E. Azhar, T. A. Madani, F. Ntoumi, R. Kock, O. Dar, G. Ippolito, T. D. McHugh, Z.A. Memish, C. Drosten, A. Zumla and E. Petersen (2020). “The continuing 2019-nCoV epidemic threat of novel coronaviruses to global health — The latest 2019 novel coronavirus outbreak in Wuhan, China.” International Journal of Infectious Diseases 91: 264–266.

Imai, N., A. Cori, I. Dorigatti, M. Baguelin, C. Donnelly, S. Riley and N. M. Ferguson (2020). Report 3: Transmissibility of 2019-nCoV London, UK, Imperial College London.

Imai, N., I. Dorigatti, A. Cori, C. Donnelly, S. Riley and N. M. Ferguson (2020). Report 2: Estimating the potential total number of novel Coronavirus cases in Wuhan City, China London, UK, Imperial College London.

Jowett, D., J. A. Browning and B. C. Haning (1974). NONLINEAR DISEASE PROGRESS CURVES.

Koopman, J. (2004). “Modeling infection transmission.” Annual Review of Public Health 25: 303–326.

Li, Q., X. Guan, P. Wu, X. Wang, L. Zhou, Y. Tong, R. Ren, K. S. M. Leung, E. H. Y. Lau, J. Y. Wong, X. Xing, N. Xiang, Y. Wu, C. Li, Q. Chen, D. Li, T. Liu, J. Zhao, M. Liu, W. Tu, C. Chen, L. Jin, R. Yang, Q. Wang, S. Zhou, R. Wang, H. Liu, Y. Luo, Y. Liu, G. Shao, H. Li, Z. Tao, Y. Yang, Z. Deng, B. Liu, Z. Ma, Y. Zhang, G. Shi, T. T. Y. Lam, J. T. Wu, G. F. Gao, B.J. Cowling, B. Yang, G. M. Leung and Z. Feng (2020). “Early Transmission Dynamics in Wuhan, China, of Novel Coronavirus–Infected Pneumonia.” New England Journal of Medicine.

Linton, N. M., T. Kobayashi, Y. Yang, K. Hayashi, A. R. Akhmetzhanov, S.-m. Jung, B. Yuan,R. Kinoshita and H. Nishiura (2020) “Epidemiological characteristics of novel coronavirus infection: A statistical analysis of publicly available case data.” medRxiv, 2020.2001.2026.20018754 DOI: 10.1101/2020.01.26.20018754.

Liu, T., J. Hu, M. Kang, L. Lin, H. Zhong, J. Xiao, G. He, T. Song, Q. Huang, Z. Rong, A. Deng,W. Zeng, X. Tan, S. Zeng, Z. Zhu, J. Li, D. Wan, J. Lu, H. Deng, J. He and W. Ma (2020) “Transmission dynamics of 2019 novel coronavirus (2019-nCoV).” bioRxiv, 2020.2001.2025.919787 DOI: 10.1101/2020.01.25.919787.

Liu, X., G. J. D. Hewings, S. Wang, M. Qin, X. Xiang, S. Zheng and X. Li (2020) “Modeling the situation of COVID-19 and effects of different containment strategies in China with dynamic differential equations and parameters estimation.” medRxiv, 2020.2003.2009.20033498 DOI: 10.1101/2020.03.09.20033498.

Mansfield, E. and C. Hensley (1960). “THE LOGISTIC PROCESS - TABLES OF THESTOCHASTIC EPIDEMIC CURVE AND APPLICATIONS.” Journal of the Royal Statistical Society Series B-Statistical Methodology 22(2): 332–337.

NHC. (2020). “National Health Commission of the People’s Republic of China.” Retrieved Feb. 8, 2020, 2020, from www.nhc.gov.cn/xcs/xxgzbd/gzbd_index.shtml.

Nishiura, H., S.-m. Jung, N. M. Linton, R. Kinoshita, Y. Yang, K. Hayashi, T. Kobayashi, B. Yuan and A. R. Akhmetzhanov (2020). “The Extent of Transmission of Novel Coronavirus in Wuhan, China, 2020.” Journal of Clinical Medicine 9(2): 330.

Roosa, K., Y. Lee, R. Luo, A. Kirpich, R. Rothenberg, J. M. Hyman, P. Yan and G. Chowell (2020). “Short-term Forecasts of the COVID-19 Epidemic in Guangdong and Zhejiang, China: February 13-23, 2020.” Journal of Clinical Medicine 9(2).

Shen, M., Z. Peng, Y. Xiao and L. Zhang (2020) “Modelling the epidemic trend of the 2019 novel coronavirus outbreak in China.” bioRxiv, 2020.2001.2023.916726 DOI: 10.1101/2020.01.23.916726.

Waggoner, P. E. and D. E. Aylor (2000). “Epidemiology: A science of patterns.” Annual Review of Phytopathology 38: 71-+.

Wang, C., P. W. Horby, F. G. Hayden and G. F. Gao (2020). “A novel coronavirus outbreak of global health concern.” The Lancet.

Wu, J. T., K. Leung and G. M. Leung (2020). “Nowcasting and forecasting the potential domestic and international spread of the 2019-nCoV outbreak originating in Wuhan, China: a modelling study.” The Lancet.

Wu, P., X. Hao, E. H. Y. Lau, J. Y. Wong, K. S. M. Leung, J. T. Wu, B. J. Cowling and G. M. Leung (2020). “Real-time tentative assessment of the epidemiological characteristics of novel coronavirus infections in Wuhan, China, as at 22 January 2020.” Eurosurveillance 25(3): 2000044.

Wu, Z. and J. M. McGoogan (2020). “Characteristics of and Important Lessons From the Coronavirus Disease 2019 (COVID-19) Outbreak in China: Summary of a Report of 72?314 Cases From the Chinese Center for Disease Control and Prevention.” JAMA.

Zhang, C. and M. Wang (2020) “MRCA time and epidemic dynamics of the 2019 novel coronavirus.” bioRxiv, 2020.2001.2025.919688 DOI: 10.1101/2020.01.25.919688.

Zhang, C. and M. Wang (2020) “Origin time and epidemic dynamics of the 2019 novel coronavirus.” bioRxiv, 2020.2001.2025.919688 DOI: 10.1101/2020.01.25.919688.

Zhao, S., Q. Lin, J. Ran, S. S. Musa, G. Yang, W. Wang, Y. Lou, D. Gao, L. Yang, D. He and M.H. Wang (2020) “Preliminary estimation of the basic reproduction number of novel coronavirus (2019-nCoV) in China, from 2019 to 2020: A data-driven analysis in the early phase of the outbreak.” bioRxiv, 2020.2001.2023.916395 DOI: 10.1101/2020.01.23.916395.

Zhao, S., Q. Lin, J. Ran, S. S. Musa, G. Yang, W. Wang, Y. Lou, D. Gao, L. Yang, D. He and M.H. Wang (2020). “Preliminary estimation of the basic reproduction number of novel coronavirus (2019-nCoV) in China, from 2019 to 2020: A data-driven analysis in the early phase of the outbreak.” International Journal of Infectious Diseases.

Zhao, S., S. S. Musa, Q. Y. Lin, J. J. Ran, G. P. Yang, W. M. Wang, Y. J. Lou, L. Yang, D. Z. Gao, D. H. He and M. H. Wang (2020). “Estimating the Unreported Number of Novel Coronavirus (2019-nCoV) Cases in China in the First Half of January 2020: A Data-Driven Modelling Analysis of the Early Outbreak.” Journal of Clinical Medicine 9(2).

Zhou, X., N. Hong, Y. Ma, J. He, H. Jiang, C. Liu, G. Shan, L. Su, W. Zhu and Y. Long (2020) “Forecasting the Worldwide Spread of COVID-19 based on Logistic Model and SEIR Model.” medRxiv, 2020.2003.2026.20044289 DOI: 10.1101/2020.03.26.20044289.

Ziff, A. L. and R. M. Ziff (2020) “Fractal kinetics of COVID-19 pandemic.” medRxiv, 2020.2002.2016.20023820 DOI: 10.1101/2020.02.16.20023820.

